# Parents’ Awareness of, Views on and Experiences with the Interim Canada Dental Benefit

**DOI:** 10.64898/2026.02.05.26345648

**Authors:** Thiago H. L. Baltus, Carol Youssef, Saif Goubran, Gurpreet Mauli, Dhaven Patel, Silvana Fux, Betty-Anne Mittermuller, Daniella DeMaré, Anil Menon, Katherine Yerex, Khalida Hai-Santiago, Sonica Singhal, Robert J. Schroth

## Abstract

**Introduction:** The Interim Canada Dental Benefit (CDB), a precursor to the Canadian Dental Care Plan (CDCP), provided financial support to low-income families of children < 12 years of age to address affordability challenges in accessing dental care. This study investigated Manitoba parents’ awareness of, views on, and experiences with the Interim CDB.

**Methods:** An interview-led survey was conducted between May through December 2023 with parents recruited through community dental clinics in Manitoba. The questionnaire captured participant demographics, awareness, views, and other experiences with the Interim CDB. Statistical analyses included descriptive statistics (frequencies, means, standard deviation), bivariate analysis (Chi-square), and logistic regression modelling. A *p*-value ≤0.05 was significant.

**Results:** Overall, 150 parents participated, with the majority being mothers (72.7%), married (69.8%), and living in urban areas (92.6%). Most parents (86.7%) had heard of the Interim CDB, but only 52.7% applied for their children, as only 48% were aware of the income criteria. Most parents (97.3%) agreed that the benefit improved access to care. Multivariate logistic regression models showed that uninsured parents were significantly more likely to have heard of the Interim CDB and to have applied for it.

**Conclusion:** Parents believed that the Interim CDB improved access to dental care. However, informational barriers remained. These findings underscore the need for simplified and inclusive communication to strengthen the reach and effectiveness of future public dental programs.

## Introduction

Oral health is a critical component of overall well-being, yet many children in Canada continue to face barriers in accessing timely and affordable dental care. Although medical services are covered under Canada’s national Medicare program, oral health care has historically remained outside the scope of universal coverage.^1^ As a result, access to dental services often depends on out-of-pocket payments or the availability of private or targeted public insurance programs.^1^ These structural limitations contribute to unmet oral health needs and delayed treatment particularly among children from low-income families, Indigenous communities, newcomers, and those living in rural and remote areas.^2^ National data show that rates of dental surgery to treat severe decay are highest among children from disadvantaged backgrounds, underscoring the scale of unmet need.^3^ Affordability is one of several key dimensions of access to oral health care, alongside availability, accommodation, acceptability, all of which influence whether families are able to obtain timely dental services for their children. ^2^

To address the affordability of dental care that contributes to these pediatric oral health disparities, the Canadian federal government introduced the Interim Canada Dental Benefit (CDB) in October 2022 as a temporary measure for children under 12 years, from families with low adjusted family net income (AFNI) and no private dental insurance.^4^ The program provided financial support for out-of-pocket dental expenses, and it was the predecessor of the Canadian Dental Care Plan (CDCP). Eligible families with annual incomes below $90,000 could receive between $260 and $650 per child, depending on income level. ^5, 6^ The benefit excluded children with private dental insurance but allowed continued eligibility for those receiving government dental assistance, provided expenses were not fully reimbursed.^7^ Applications required prior-year tax filing, and payments were administered through the Canada Revenue Agency (CRA).^6, 7^ Period 1 covered treatment from October 1^st^, 2022, to June 30^th^, 2023, and Period 2 from July 1^st^ of 2023, to June 30^th^ of 2024.^7^

To date, studies exploring families’ experiences with the Interim CDB and whether it improved access to dental care for their children are lacking. Understanding parents’ perspectives is essential for assessing how temporary benefits function in practice and for informing the rollout of broader public dental programs like the CDCP. The purpose of this study was to investigate parents’ awareness, views, and experiences with the Interim CDB.

## Methods

This study was approved by the University of Manitoba’s Health Research Ethics Board (HS26028/H2023:180). The questionnaire was developed by the study team, including Health Canada’s Oral Health Branch staff, and was informed by questions included in a survey conducted by the Strategic Counsel, through a signed contract with the Health Canada’s Oral Health Branch^8^. The questionnaire contained 48 questions, both open and closed-ended, intended to investigate the respondents’ characteristics and views. The questionnaire included seven sections: 1) demographic characteristics; 2) awareness of the Interim CDB; 3) views on the application process; 4) information about receiving the Interim CDB; 5) information about utilizing the Interim CDB; 6) perceived challenges with the Interim CDB; and 7) suggestions for program improvement.

Parents and caregivers (referred to as “parents” in the remainder of this manuscript) residing in Manitoba were eligible to participate if they had a child under 12 years who received care at a community-based public dental clinic or had previously provided consent to be contacted for research. The study relied on a convenience sampling method for participant recruitment.

The questionnaire was administered from May 2023 through December 2023. Data were collected using a structured, interviewer-administered questionnaire created in Microsoft Forms. The area of residence was determined as urban or rural based on the postal code provided by the participant. Interviews were conducted in English and professional language interpreters with Shared Health’s Language Access Program were used when necessary for those with language barriers. Interviews were conducted in person, by phone, or via video call (e.g., Zoom, Microsoft Teams), depending on the participant’s preference and accessibility. Informed consent was obtained in writing from all participants interviewed in person while verbal (audio-recorded) consent was obtained using a standardized script for surveys conducted via phone or video call.

Data were exported from Microsoft Forms to Excel (Microsoft Inc., USA) and analyzed using Number Cruncher Statistical Software (NCSS Version 24.0.2; Kaysville, Utah). Descriptive statistics included frequencies, means, and standard deviations (SD). Bivariate analyses included Chi-squared and Fisher’s exact tests, while evaluating the outcomes “heard about the Interim CDB” and “applied for the Interim CDB”. Multivariate logistic regression analysis was performed, and two models were created. The first model used the outcome “heard about the Interim CDB”, while the second model used the outcome “applied for the Interim CDB”. For each model, covariates were selected based on the associations observed in the bivariate analysis and those having an association with the respective outcomes of a p value <0.55 included. Results from multivariate logistic regression were reported as regression coefficients, odds ratios (OR), and 95% confidence intervals (95% CI). For all tests, *p*-values ≤0.05 were considered significant.

## Results

Overall, 150 parents were enrolled, the majority of whom were mothers (72.7%), married (69.8%), employed (53.1%), living in urban areas (92.6%), and having 2.8 ± 1.5 children (Table 1). Most respondents indicated that they lacked dental insurance (62.0%). Among those with dental insurance, one-third (33.3%) had an employer-sponsored plan while 31.6% had Non-Insured Health Benefits (NIHB) through the Department of Indigenous Services Canada, and 27.8% had Employment and Income Assistance (EIA).

**Table 1:** Demographic and Socioeconomic Characteristics.

| Variable | n (%) total = 150 |
| --- | --- |
| Relationship to child (n=150) |  |
| Mother | 109 (72.7) |
| Father | 32 (21.3) |
| Grandparent | 7 (4.7) |
| Other | 2 (1.3) |
| Area (n=148) |  |
| Urban | 137 (92.6) |
| Rural | 11 (7.4) |
| Number of children* | 2.8 ($\pm 1.5$) |
| Group parent self-identify with (n=149) |  |
| African | 48 (32.2) |
| Asian | 44 (29.5) |
| Canadian Indigenous | 29 (19.5) |
| Caucasian | 23 (15.4) |
| Other | 5 (3.4) |
| Parent marital status (n=149) |  |
| Married | 104 (69.8) |
| Single | 37 (24.8) |
| Living with partner | 5 (3.4) |
| Other | 3 (2.0) |
| Parental education level (n=148) |  |
| Lower than High School | 23 (15.5) |
| High School diploma | 41 (27.7) |
| Higher than High School | 84 (56.8) |
| Employment status (n=147) |  |
| Yes, full-time | 46 (31.3) |
| Yes, part-time | 32 (21.8) |
| No | 69 (46.9) |
| Insurance or government program that covers for dental (n=150) |  |
| Has insurance | 57 (38.0) |
| Does not have insurance | 93 (62.0) |
| Type of insurance or program (n=57) |  |
| An employer-sponsored plan | 17 (29.8) |
| Non-Insured Health Benefits (NIHB) | 16 (28.1) |
| Employment and Income Assistance (EIA) | 15 (26.3) |
| Private insurance plan | 5 (8.8) |
| Other | 4 (7.0) |

\*mean ( ± standard deviation)

Responses to questions about awareness of and the application process for the Interim CDB are reported in Table 2. The majority of participants had heard of the Interim CDB (86.7%), with most learning about the program from dental offices (27.9%), family or friends (17.0%), and the CRA website (13.6%). Despite the high awareness of the program, many were unaware of the eligibility criteria. For instance, only 59.3% knew that those with private dental insurance were ineligible for the Interim CDB, while 60.7% knew that receiving the Canada Child Benefit was an eligibility requirement. Many respondents were also unaware that children < 12 years of age were eligible even if their families had public dental insurance, such as the NIHB program (65.3%) and provincial EIA programs (59.1%). Additionally, nearly half of participants (48.0%) reported knowing that there were specific income cutoff thresholds for eligibility, while 44.0% knew about the possibility of applying for the Interim CDB after they had already paid out of pocket for their child’s dental care.

**Table 2:** Awareness of Interim CDB and Application procedure.

| Variable | n (%) total = 150 |
| --- | --- |
| Heard about the Interim CDB (n=150) |  |
| Yes | 130 (86.7) |
| No | 20 (13.3) |
| Knew that receiving the CCB<sup>a</sup> was an eligibility requirement to receive the Interim CDB (n=150) |  |
| Yes | 91 (60.7) |
| No | 59 (39.3) |
| Knew that children with private dental insurance were not eligible to receive the Interim CDB (n=150) |  |
| Yes | 89 (59.3) |
| No | 61 (40.7) |
| Knew that families that received EIA<sup>b</sup> were eligible to receive the Interim CDB (n=149) |  |
| Yes | 61 (40.9) |
| No | 88 (59.1) |
| Knew that families that received NIHB<sup>c</sup> were eligible to receive the Interim CDB (n=150) |  |
| Yes | 52 (34.7) |
| No | 98 (65.3) |
| Knew that parents had to be Canadian citizens or Permanent Residents of Canada to be eligible to receive the Interim CDB (n=150) |  |
| Yes | 107 (71.3) |
| No | 43 (28.7) |
| Was aware of the income cut-offs to receive the Interim CDB (n=150) |  |
| Yes | 72 (48.0) |
| No | 78 (52.0) |
| Received enough information about the Interim CDB (n=150) |  |
| Strongly agree | 11 (7.3) |
| Agree | 66 (44.0) |
| Neutral | 24 (16.0) |
| Disagree | 39 (26.0) |
| Strongly disagree | 10 (6.7) |
| Visited the Government of Canada website for more information about the Interim CDB (n=150) |  |
| Yes | 75 (50.0) |
| No | 75 (50.0) |
| Thought that the information provided by the Government about the Interim CDB was clear (n=150) |  |
| Yes | 89 (59.3) |
| No | 34 (22.7) |
| Not sure | 27 (18.0) |
| Learned about the Interim CDB from (n=147) |  |
| Dental Office | 41 (27.9) |
| Family/Friends | 25 (17.0) |
| Government website (CRA<sup>d</sup>) | 20 (13.6) |
| Social media | 15 (10.2) |
| Government letter (CRA<sup>d</sup>) | 14 (9.5) |
| Members of this study team | 13 (8.8) |
| TV/Radio | 11 (7.5) |
| Community group | 2 (1.4) |
| Other | 5 (3.4) |
| Not aware | 1 (0.7) |
| Knew to keep the children's dental treatment receipts after using the Interim CDB (n=150) |  |
| Yes | 58 (38.7) |
| No | 92 (61.3) |
| Knew that they could apply for the Interim CDB after child received dental treatment (n=150) |  |
| Yes | 66 (44.0) |
| No | 84 (56.0) |
| Applied for the Interim CDB (n=148) |  |
| Yes | 79 (53.4) |
| No | 69 (46.6) |
| Felt uncomfortable with the questions (for those who applied) (n=76) |  |
| Yes | 3 (3.9) |
| No | 73 (96.1) |
| Reasons why people did not apply for the benefit (n=69) |  |
| Did not know about the program | 18 (25.6) |
| Did not meet eligibility criteria | 12 (17.9) |
| Have private insurance | 10 (14.1) |
| No time to apply | 6 (9.0) |
| Not receiving CCB<sup>a</sup> | 3 (3.8) |
| Child over 12 | 3 (3.8) |
| Language barrier | 2 (2.6) |
| Did not file income taxes in the past two years | 2 (2.6) |
| No one to help with application | 2 (2.6) |
| Technology barriers (no phone, computer, or access to Wi-Fi) | 1 (1.3) |
| Other | 10 (16.7) |
| Method of application for the Interim CDB (n=77) |  |
| CRA<sup>d</sup> website | 46 (59.7) |
| By phone | 30 (39.0) |
| QR code | 1 (1.3) |
| Difficulty of the overall application process (n=78) |  |
| Hard | 5 (6.4) |
| Moderate | 23 (29.5) |
| Easy | 50 (64.1) |
| Met challenges while applying for the Interim CDB (n=77) |  |
| Yes | 16 (20.8) |
| No | 61 (79.2) |
| Received a letter from the CRA<sup>d</sup> about the Interim CDB (n=139) |  |
| Yes | 47 (33.8) |
| No | 92 (66.2) |
| Was confused by the CRA<sup>d</sup> letter (for those who received it) (n=50) |  |
| Yes | 9 (18.0) |
| No | 41 (82.0) |
| Required language/translation assistance to apply for the Interim CDB (n=82) |  |
| Yes | 24 (29.3) |
| No | 58 (70.7) |
| Knew that could ask the CRA<sup>d</sup> for an interpreter to provide language support the application (n=145) |  |
| Yes | 41 (28.3) |
| No | 104 (71.7) |
| Were aware of the possibility of reapplying in July 2023 (n=141) |  |
| Yes | 57 (40.4) |
| No | 84 (59.6) |

<sup>a</sup>CCB = Canada Child Benefit
<sup>b</sup>EIA = Employment and Income Assistance
<sup>c</sup>NIHB = Non-Insured Health Benefits
<sup>d</sup>CRA = Canada Revenue Agency

Overall, only 53.4% participants applied for the Interim CDB (Table 2). Most applicants (59.7%) applied online through the CRA website, while 39.0% applied by phone. A majority (64.1%) described the application process as easy, while 20.8% reported challenges. Of those who applied, 29.3% required language interpreter assistance when applying. The most common reasons for not applying included being unaware of the program (25.6%), not meeting eligibility criteria (17.9%), and having access to private dental insurance for their children (14.1%). Unfortunately, only 40.4% of the entire survey sample were aware of the ability to reapply for the second year of the Interim CDB program.

Responses to questions about the receipt and use of the Interim CDB are presented in Table 3. Half (50.4%) of participants reported receiving the Interim CDB for their children. Nearly all (95.5%) who received the benefit reported receiving the full amount per child ($650). Most recipients (85.9%) agreed that the amount was sufficient to cover their child’s dental expenses. Overall, 97.3% of parents believed that the Interim CDB improved access to oral health care for young children. However, 41.4% of respondents were dissatisfied or very dissatisfied that the Interim CDB provided assistance only for children ≤ 12 years of age. Apart from affordability, some common barriers to accessing dental care reported by respondents included transportation (13.1%), language barriers (9.9%), lack of family support (7.2%), and dental offices operating only during daytime hours (7.2%). Participants were also asked to provide feedback and suggestions they had to improve the Interim CDB (Table 3). Respondents indicated that information about the program should be available on social media (20.8%), in dental offices (18.5%), in schools and daycare facilities (18.0%), and in physicians’ offices (15.7%). Fortunately, the majority (80.1%) indicated that resources about the Interim CDB were in a language they could understand, and that staff from dental offices were helpful and knowledgeable about the program (72.5%). Nearly every respondent (99.3%) indicated that the Interim Canada Dental Benefit should have included children up to the age of 18 years.

**Table 3:** Answers for Receiving, Using, Challenges with and Suggestion for the Interim CDB.

| Variable | n (%) total = 150 |
| --- | --- |
| Received the Interim CDB (n=136) |  |
| Yes | 69 (50.7) |
| No (or N/A)<sup>a</sup> | 67 (49.3) |
| Reasons why the Interim CDB was denied (n=24) |  |
| Have private insurance | 10 (41.6) |
| Not receiving Canada Child Benefit | 4 (16.7) |
| Did not file income taxes in the past two years | 4 (16.7) |
| Child over 12 | 2 (8.3) |
| Earning more than \$90,000 | 1 (4.2) |
| Other | 3 (12.5) |
| Amount received per child (n=66) |  |
| \$650 | 63 (95.5) |
| \$390 | 3 (4.5) |
| \$260 | 0 |
| The amount received covered the dental costs for treatment (n=64) |  |
| Yes | 55 (85.9) |
| No | 9 (14.1) |
| Time until the benefit was received (n=67) |  |
| Within one week | 45 (67.1) |
| Within 2-3 weeks | 20 (30.0) |
| Within 3-4 weeks | 2 (2.9) |
| Child has been to the dentist before receiving the Interim CDB (n=143) |  |
| Yes | 123 (86.0) |
| No | 20 (14.0) |
| Believed that their child needed urgent dental care (n=148) |  |
| Yes | 59 (39.9) |
| No | 89 (60.1) |
| Would have taken their child to the dentist even if had not received the Interim CDB (n=145) |  |
| Yes | 126 (86.9) |
| No | 19 (13.1) |
| Believed that the Interim CDB improved access to oral health care for children (n=146) |  |
| Yes | 142 (97.3) |
| No | 4 (2.7) |
| Reasons that make accessing dental care difficult (n=150) |  |
| No barriers | 40 (27.0) |
| Transportation | 20 (13.1) |
| Language barrier | 15 (9.9) |
| No support | 11 (7.2) |
| Dental office closes after the workday | 11 (7.2) |
| Cannot take time off work | 10 (6.7) |
| Single parent | 10 (6.7) |
| Finding daycares for other children | 9 (6.3) |
| Technology barriers (no phone, computer, or access to Wi-Fi) | 7 (4.8) |
| No dental services available in my area | 4 (2.8) |
| Other | 13 (8.3) |
| Likelihood of taking their child to the dentist after receiving the Interim CDB (n=137) |  |
| Very likely | 76 (55.5) |
| Likely | 34 (24.8) |
| Neutral | 24 (17.5) |
| Unlikely | 3 (2.2) |
| Satisfaction over the Interim CDB age coverage (only for children ≤12 years) (n=150) |  |
| Very Satisfied | 20 (13.3) |
| Satisfied | 36 (24.0) |
| Neutral | 32 (21.3) |
| Dissatisfied | 42 (28.1) |
| Very Dissatisfied | 20 (13.3) |
| Applied for the Interim CDB additional payment when treatment costs exceeded the amount received (n=76) |  |
| Yes | 10 (13.2) |
| No | 42 (55.3) |
| Not aware this was an option | 24 (31.5) |
| Considered saving the funds from the Interim CDB to pay for more expensive dental treatments (e.g., root canals, braces, etc.) (n=77) |  |
| Yes | 47 (61.0) |
| No | 30 (39.0) |
| Thought that more information on the Interim CDB should be found at (n=150) |  |
| Social media | 32 (20.8) |
| Dental offices | 28 (18.5) |
| Schools/Daycares | 27 (18.0) |
| Doctor's offices | 23 (15.7) |
| Community centers/fairs | 22 (14.6) |
| Community newspapers | 15 (10.2) |
| Other | 3 (2.2) |
| The resources for the Interim CDB were in a language they could understand (n=146) |  |
| Yes | 117 (80.1) |
| No | 28 (19.2) |
| Don't know | 1 (0.7) |
| Staff at the dental office/clinic were helpful/knowledgeable about the Interim CDB (n=149) |  |
| Yes | 108 (72.5) |
| No | 22 (14.8) |
| N/A | 19 (12.7) |
| Believed that the benefit should cover children up to 18 years of age (n=150) |  |
| Yes | 149 (99.3) |
| No | 1 (0.7) |

<sup>a</sup> Respondents who did not apply were included as 'not applicable' for not receiving the benefit.

Two key variables of interest in this study included whether participants had heard about the Interim CDB and whether they had applied for the program for their children. Results from the bivariate analysis of participant characteristics and these two outcomes are presented in Table 4. Overall, participants who self-declared Asian were statistically the most likely (76.7%) to have applied for the Interim CDB, while self-declared Caucasians were the least likely to have applied (26.1%). In both outcomes, the dental insurance situation showed statistically significant differences. A significant association between the outcomes “Heard about the Interim CDB” and “Applied for the Interim CDB” was observed (p<0.001).

**Table 4.**
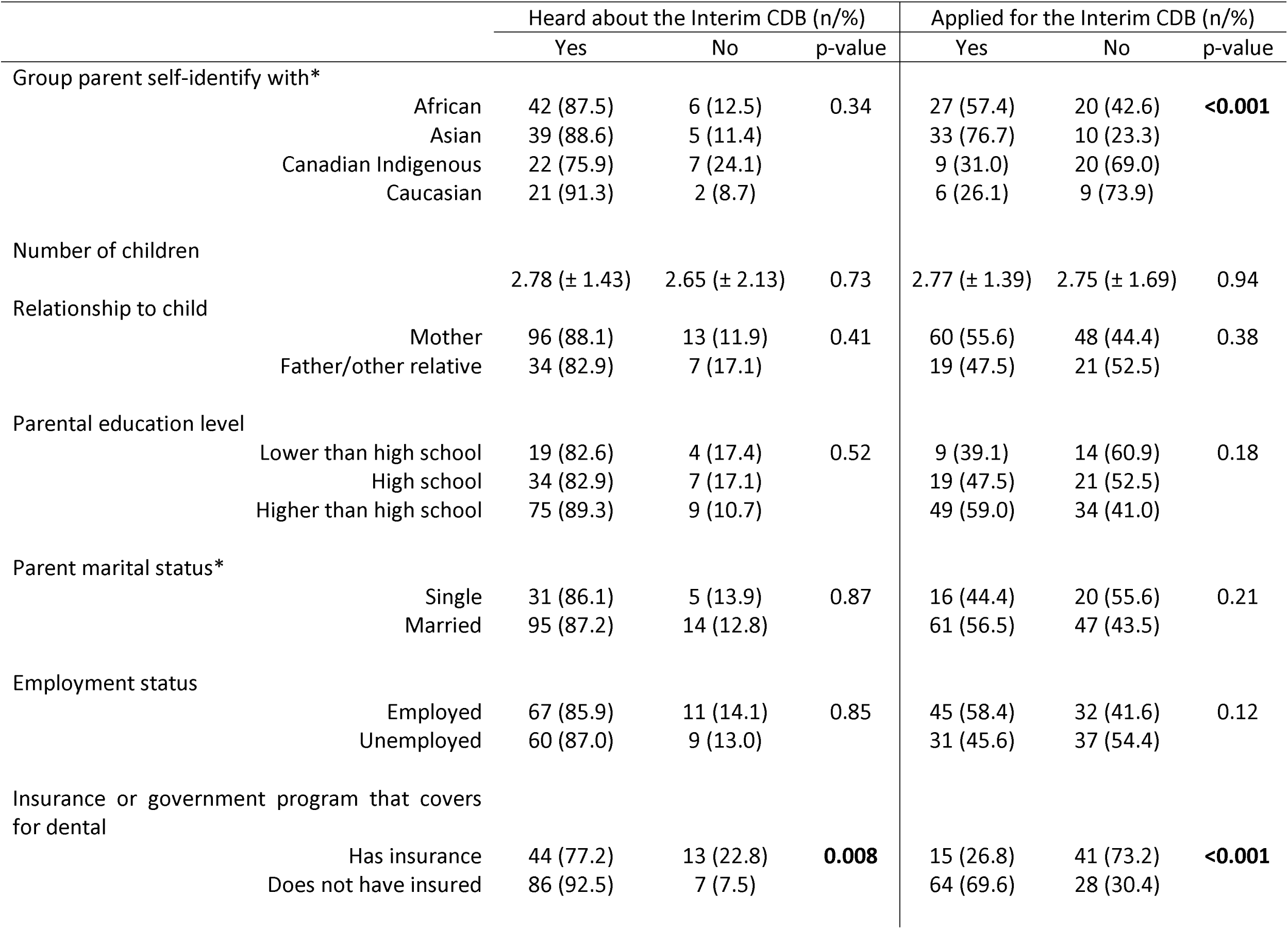
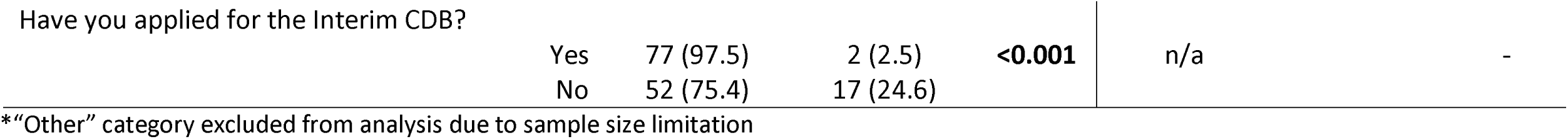
Correlations between participant’s sociodemographic characteristics and survey questions.

Lastly, the results from the multivariate logistic regression models are presented in Table 5. The absence of dental insurance was significantly associated with the two proposed outcomes: (model one first - model two later) uninsured parents were significantly more likely to have heard about (OR = 1.89, p = 0.03) and to have applied for (OR = 2.37, p<0.001) the Interim CDB, in comparison to parents with dental coverage. For Model 2, when using African (self-identified) parents as reference for comparison, Asian participants were more likely to have applied for the benefit (OR = 3.05, p = 0.003), while Caucasian parents were less inclined to apply (OR = 0.39, p = 0.02).

**Table 5.** Multivariate logistic regression models for answers adjusted for demographic parameters.

| **Model 1** |  | Heard about the Interim Canada Dental Benefit<sup>a</sup> |  |  |
| --- | --- | --- | --- | --- |
| Variable | Coefficient | Odds ratio (OR) | 95% Confidence Interval for OR | p-value |
| Intercept | - 0.06 | - | - | - |
| Group self-identify with |  |  |  |  |
| Asian<sup>b</sup> | - 0.17 | 0.85 | 0.61 - 1.47 | 0.72 |
| Canadian Indigenous<sup>b</sup> | - 0.34 | 0.71 | 0.34 - 2.10 | 0.45 |
| Caucasian<sup>b</sup> | 0.60 | 1.83 | 0.55 - 6.11 | 0.33 |
| Relationship to child |  |  |  |  |
| Mother<sup>c</sup> | - 0.25 | 0.78 | 0.46 - 1.33 | 0.36 |
| Parental education level |  |  |  |  |
| High school<sup>d</sup> | - 0.40 | 0.67 | 0.31 - 1.42 | 0.30 |
| Higher than high school<sup>d</sup> | 0.31 | 1.37 | 0.67 - 2.77 | 0.39 |
| Insurance or government program that covers for dental |  |  |  |  |
| Does not have insurance<sup>e</sup> | 0.63 | 1.89 | 1.05 - 3.38 | **0.03** |
| **Model 2** |  | Applied for the Interim Canada Dental Benefit<sup>a</sup> |  |  |
| Variable | Coefficient | Odds ratio (OR) | 95% Confidence Interval for OR | p-value |
| Intercept | - 0.72 | - | - | - |
| Group self-identify with |  |  |  |  |
| Asian<sup>b</sup> | 1.12 | 3.05 | 1.44 - 6.45 | **0.003** |
| Canadian Indigenous<sup>b</sup> | - 0.25 | 0.78 | 0.32 - 1.90 | 0.58 |
| Caucasian<sup>b</sup> | - 0.93 | 0.39 | 0.17 - 0.89 | **0.02** |
| Relationship to child |  |  |  |  |
| Mother<sup>c</sup> | 0.24 | 1.27 | 0.80 - 2.01 | 0.32 |
| Parental education level |  |  |  |  |
| High school<sup>d</sup> | - 0.28 | 0.76 | 0.41 - 1.41 | 0.38 |
| Higher than high school<sup>d</sup> | 0.37 | 1.44 | 0.81 - 2.56 | 0.21 |
| Insurance or government program that covers for dental |  |  |  |  |
| Does not have insurance<sup>e</sup> | 0.86 | 2.37 | 1.48 - 3.80 | **<0.001** |
| Parent marital status |  |  |  |  |
| Single<sup>f</sup> | 0.74 | 2.10 | 0.62 - 7.09 | 0.23 |
| Other<sup>f</sup> | - 0.79 | 0.45 | 0.06 - 3.71 | 0.46 |
| Employment status |  |  |  |  |
| Employed<sup>g</sup> | 0.21 | 1.23 | 0.80 - 1.89 | 0.34 |

References: <sup>a</sup>Did not apply for the Interim Canada Dental Benefit <sup>b</sup>African <sup>c</sup>Father/other relative <sup>d</sup>Lower than high school <sup>e</sup>Has insurance <sup>f</sup>Married <sup>g</sup>Unemployed

## Discussion

This study of Manitoba parents provides a glimpse into their awareness, views and experiences with the Interim CDB. While most participants were aware of the benefit, just over half reported having applied for it for their children. This gap between awareness and application suggests that knowledge of a program alone is insufficient to ensure uptake, highlighting the importance of clarity, trust, and ease of navigation in public benefit design. This finding was somewhat surprising considering that parents were primarily recruited from clinics serving low-income families. One reason for this lower enrollment in the program despite its awareness may be attributed to parents being uncertain about eligibility criteria and how to apply for the Interim CDB. It is also possible that many parents faced barriers that prevented them from applying, including potential distrust of the CRA, language and literacy challenges, and perceived complexity of the application process

A positive finding in this study was that the majority of participants believed that the Interim CDB improved access to dental care for children < 12 years of age. Facilitating access to oral health care for children under 12 was the primary reason for establishing the Interim CDB. Affordability is a major domain of access to care, and many Canadians have avoided care because of the cost of oral health services.^9^ The introduction of the Interim CDB was the first step in the federal government’s attempt to introduce dental care support for lower income Canadians. Some of the barriers to accessing dental care reported by participants in this study included transportation and language barriers. Transportation is often reported as a barrier to accessing oral health care, particularly for people living in rural or remote areas, which is aggravated by factors such as availability of means of transport, distance, weather conditions, etc.^9, 10^ To expand the reach of the benefit and target those with language barriers, Health Canada’s Oral Health Branch developed promotional material in many languages, including Punjabi, Michif Cree, Somali, simplified and traditional Chinese, Arabic, Tagalog, Ukrainian and Spanish. ^11^

Awareness of the Interim CDB was high among this survey population. This may be because the participants were recruited from community-based dental public health clinics that may have been actively promoting the Interim CDB. Additionally, the majority of the participants had a higher level of education than what is often found in community-based studies involving low-income families. Surprisingly, a recent national survey identified that respondents from Manitoba and Saskatchewan had the lowest awareness of the Interim CDB.^9^

Regression analysis of awareness of the Interim CDB revealed that participants without dental insurance were significantly more likely to have heard about the Interim CDB after controlling for covariates such as ethnic background, relationship to the child and parent education level. This may reflect that how uninsured families were either more interested or in higher need of learning about options to support the dental care needs of their children. These findings highlight the importance of developing effective strategies to maximize reach when implementing new public policies.

The second regression model explored those factors associated with whether parents applied for the benefit. That logistic regression model reveals that ethnic background and not having insurance or a government program that covers dental were the variables significantly associated with higher odds of parents applying for the Interim CDB. This variation observed among parents from different ethnic backgrounds could potentially be an indicator of the uneven reach of the benefit’s information, despite Health Canada’s advertising strategies. These findings should be interpreted cautiously, as ethnicity may reflect intersecting factors such as information pathways, community networks, language access, and prior engagement with public programs rather than ethnicity itself. There is little surprise that those lacking insurance were more likely to have applied to the program, as affordability is a significant factor in determining access to oral health care. Interestingly, this study had a higher proportion of participants who reported having applied to the benefit than those from Manitoba and Saskatchewan (18.3%) reported by Menon et al. ^9^

Overall, opinions on aspects of the benefit were quite diverse. Only half of the participants reported having received enough information regarding the Interim CDB, and only 59.3% agreed that the information provided by the Government was clear. These communication problems could have resulted from the rapid pace at which the Interim CDB was implemented. This may explain why the official Interim CDB website was only the third most common source of information on the program that participants mentioned. Most participants indicated that they learned about the program directly from dental office staff directly, which highlights the central role oral health professionals play in disseminating information about public dental benefits. This finding positions dental offices as critical intermediaries in the successful implementation of publicly funded oral health programs. ^12^ Taken together, these findings underscore the importance of effectively disseminating information to the public and the oral health professionals.

This study is not without limitations. Data were collected through a parent survey, and results reflect the experiences of those who chose to participate and had the means to do so. As a self-reported survey, responses were subject to recall and social desirability bias. Some parents may have overstated their knowledge of the benefit or engagement with dental services. While participants were asked to reflect on their experiences with the Interim CDB, we were unable to verify the accuracy of reported dental visits, application outcomes, or how benefit funds were ultimately used. Additionally, this study was conducted in Manitoba and with a low-income, predominantly urban population. Thereby, findings may not be entirely generalizable to parents in other regions in Canada, particularly considering those living rurally. However, those participating in this study were the actual population being targeted with the Interim CDB. Since the samples were obtained by convenience, limitations on sample size should also be considered. For example, multivariate logistic models could not have been adjusted for area of residence (determined as “urban” or “rural” based on the postal code provided by the participant) due to sample size limitations.

## Conclusion

The Interim CDB was a meaningful first step toward improving access to dental care for children in lower-income families. In this study, parents in Manitoba overwhelmingly supported the benefit and reported that it helped them seek care they may not have otherwise been able to afford. At the same time, many parents faced challenges navigating the application process, understanding eligibility criteria, and accessing language or logistical support. Some families continued to delay care due to cost, transportation, or difficulty taking time off work. These findings point to the need for clearer communication, simplified processes, and inclusive outreach as the Canada Dental Care Plan (CDCP) moves forward. As the CDCP expands nationally, leveraging trusted points of contact such as dental offices and ensuring multilingual, accessible communication will be essential to translating coverage into actual care. Building on the early successes of the Interim CDB, future programs must also address the broader barriers families face to ensure equitable and consistent access to oral health care.

## Data Availability

All data produced in the present work are contained in the manuscript

## Authors contributions

### Funding

Operating funds for this study were provided by Dr. Schroth’s Canadian Institutes of Health Research Applied Public Health Chair Award in “Public Health Approaches to Improve Access to Oral Health Care and Oral Health Status for Young Children in Canada”.

## Acknowledgments

Dr. Robert J Schroth holds a Canadian Institutes of Health Research Applied Public Health Chair in “Public Health Approaches to Improve Access to Oral Health Care and Oral Health Status for Young Children in Canada”.

Dhaven Patel held an Undergraduate Research Award at the University of Manitoba

